# London Ramadan Fasting Study (LORANS): Rationale, design, and methods

**DOI:** 10.1101/2021.07.14.21260518

**Authors:** Rami Al-Jafar, Paul Elliott, Konstantinos K. Tsilidis, Abbas Dehghan

**Author notes:** Correspondence to: Abbas Dehghan, Department of Epidemiology and Biostatistics, School of Public Health, Imperial College London, 1st floor, Norfolk Place, W2 1 PG, London, UK., Rami Aljafar, Department of Epidemiology and Biostatistics, School of Public Health, Imperial College London, 1st floor, Norfolk Place, W2 1 PG, London, UK.

## Abstract

**Background:** Hundreds of millions of Muslims fast during the month of Ramadan. The London Ramadan Fasting Study (LORANS) aims to assess the lifestyle changes during this month and investigate the effect of Ramadan fasting on health.

**Methods:** LORANS is an observational study of participants that follow religious fasting in Ramadan. We advertised, recruited, and visited participants in five mosques in London, United Kingdom. In total, 146 individuals were recruited before Ramadan in May 2019 of which 85 participated in the follow up visit after Ramadan. The study protocol was approved by the ethics committee affiliated to Imperial College London. A written informed consent was signed by all the participants. Every participant completed a questionnaire, a physical examination, and gave blood samples at each visit. Moreover, they completed a 3-day food diary before Ramadan and once again during Ramadan to record dietary changes during the month of fasting.

**Results:** The mean age of participants was 45.6± 15.9 years. 47.1% of the participants were females, 25.5% were obese, 4.7% were smokers, 14% were diabetic, 24% were hypertensive, and 5.2% had cardiovascular diseases. Data collection covered demographics, lifestyle, food intake, blood pressure, anthropometric measurements, body composition, and metabolic biomarker profiling.

**Conclusion:** By engaging with mosques, proper introduction of the study aims and convenient recruitment in the mosque, we were able to recruit a balanced population regarding age and sex and collected valuable data on Ramadan fasting using high-quality techniques.

## Introduction

Ramadan is the ninth month of the Islamic calendar. During Ramadan, Muslims fast from dawn until dusk. During this time, food and drinks are prohibited at least 11 hours a day for 28–30 days (1). Fasting is one of the five pillars of Islam that all Muslims must perform with few exceptions such as children, patients and travellers (2). Fasting brings in a major change in the lifestyle, including change in dietary intake, dietary diversity and dietary habits as well as drinking, smoking, physical activity, and even sleep patterns. Since over a billion Muslims might practice fasting every year, assessing its health effects is of clinical and public health importance.

Despite the widespread practice of Ramadan fasting, high-quality research is scarce on this area. Few studies have assessed the effect of Ramadan fasting on certain cardiometabolic risk factors, including weight, blood pressure, levels of serum glucose, lipids and nutritional intake. In most instances, the studies have reported contradicting conclusions. For instance, several researchers (3-11) reported a significant decrease in weight, BMI and waist circumference (WC) after fasting during Ramadan. No changes were noted on these parameters in some other studies (12-21). Blood pressure is another important risk factor for chronic diseases and has been reported to decrease following Ramadan fasting in some studies (22-26), but not to be influenced in some others (12, 27-30). Serum glucose was also reported to drop after Ramadan fasting in two studies (4, 31) and was higher in a separate study (8). Similarly, studies are not conclusive on how serum levels of HDL and LDL cholesterol are affected by Ramadan fasting. Some studies have shown a decrease in LDL and an increase in HDL (32), while others found opposite (7, 33) or no association (8). The heterogeneity in the findings and lack of conclusiveness might be due to the limitations of these studies. Most of these studies are very small (sample sizes even less than 25), have recruited participants from only one ethnic group, and applied convenience sampling that does not represent the whole population (e.g. students). Such limitations may affect the validity of the results and hamper their generalizability (34, 35).

Moreover, the effects of Ramadan fasting might be variable in reality due to the wide range of variations in traditions attached to fasting, differences in cultures and various environmental factors. Altitude, for instance, is an essential factor that determines the duration of fasting. The exact number of hours that people should fast during Ramadan varies based on the altitude of the place but is generally between 11 to 18 hours (36). Moreover, working hours are reduced during Ramadan in some Islamic countries such as UAE, Saudi Arabia, Egypt, and Malaysia (37), while it stays the same in many other countries. Finally, socioeconomic status, level of physical activity, smoking, and alcohol consumption might modify the effect of Ramadan fasting (38).

Altogether, there is a need for large scale high-quality research that covers different age groups and various ethnicities to highlight the health effects of fasting in Ramadan on health. London is a multi-ethnic metropolitan city where approximately 1 million Muslims live. This allows a comprehensive study on Ramadan fasting including various ethnicities and cultures. Therefore, we conducted the London Ramadan Fasting Study (LORANS) to investigate the effect of Ramadan fasting on blood pressure, anthropometry/body composition, food intake and metabolic biomarkers.

## Methods

LORANS is an observational study consisting of two visits (before and after Ramadan) that was conducted to evaluate the impact of Ramadan fasting on health. Initially, we contacted six large mosques in London and invited them to support our study. Five large mosques including Almanaar Mosque (https://almanaar.org.uk/), Beitulfutuh Mosque (www.baitulfutuh.org), Madina Mosque (https://madina-masjid.org.uk/), Finsbury Park Mosque (http://www.finsburyparkmosque.org/), and West London Mosque (https://almuntadatrust.org/) agreed to collaborate with us (**Fig. 1**). We advertised the study via the mosques’ social media channels (Facebook page, Twitter account and newsletters). The announcements offered the opportunity to participate in a study exploring the impact of Ramadan fasting on health.

**Figure1:**
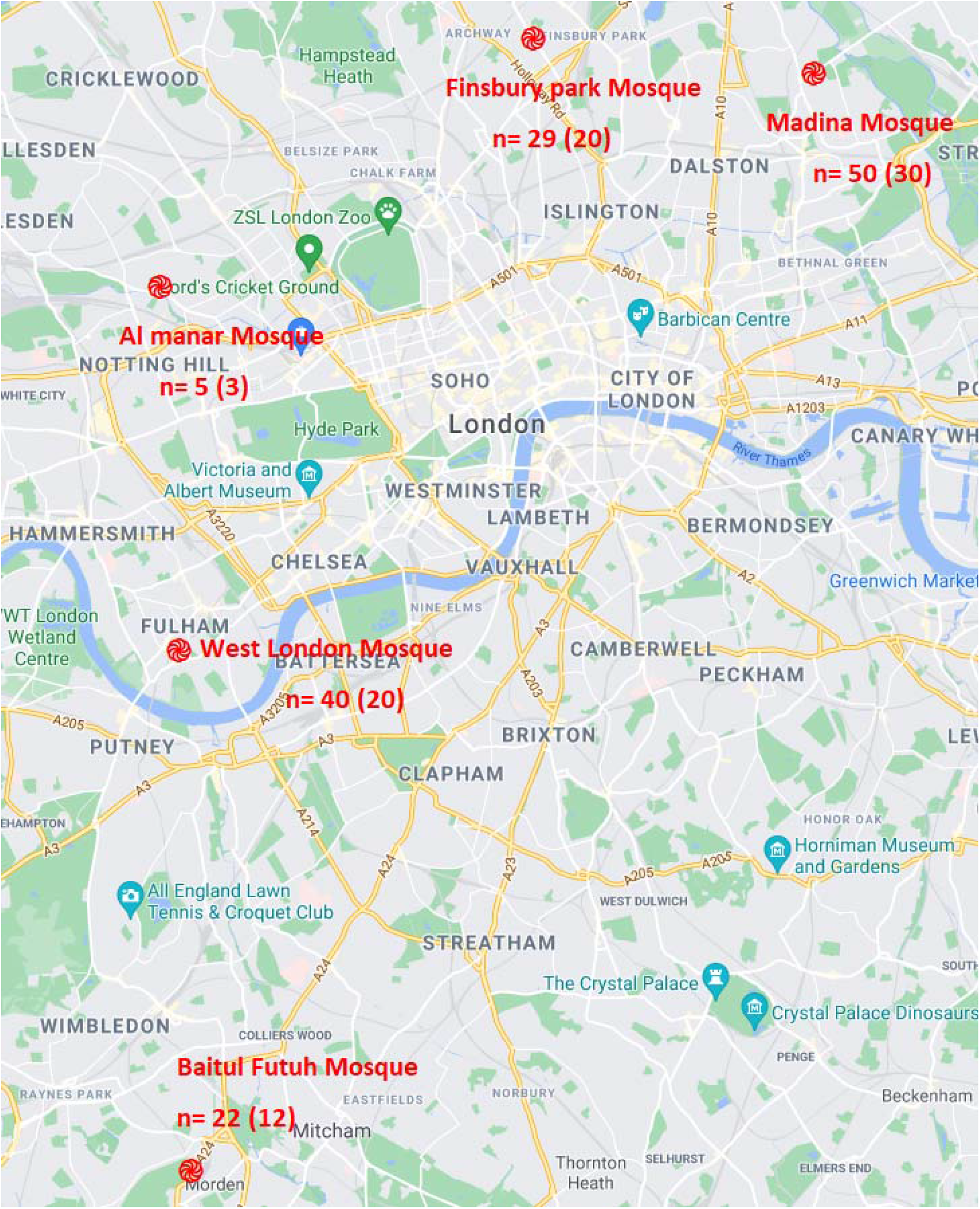
Mosques locations and numbers as baseline (follow up) by mosques.

Moreover, we spoke to the public in the mosques during daily and Friday prayers. We installed several posters and our research assistants distributed invitation flyers in the mosques. Those who were willing to participate were able to make an appointment for the first visit via the study website or a phone number. Also, we allowed individuals without appointments to walk in and take part in the study on the recruitment days. Each participant received the results of physical examinations and blood tests for the tests before and after Ramadan, which could indicate the impact of Ramadan fasting on their health.

### Inclusion and exclusion criteria

An individual had to be 18 years or older and planning to fast at least 20 days of Ramadan to participate in the study. We excluded pregnant women and individuals who were not able to attend the second visit after Ramadan.

Measurements and data were collected from every participant twice, before Ramadan (26 April – 30 April) and after Ramadan (10 June – 14 June) (**Fig. 2**). The examinations were done by three teams that rotated across the mosques. Each team consisted of a registered phlebotomist and two research assistants (male and female). The teams were trained in two sessions in advance of the study in a clinic similar to those at mosques to learn the protocols and harmonise the measurements and data collection.

**Figure 2:**
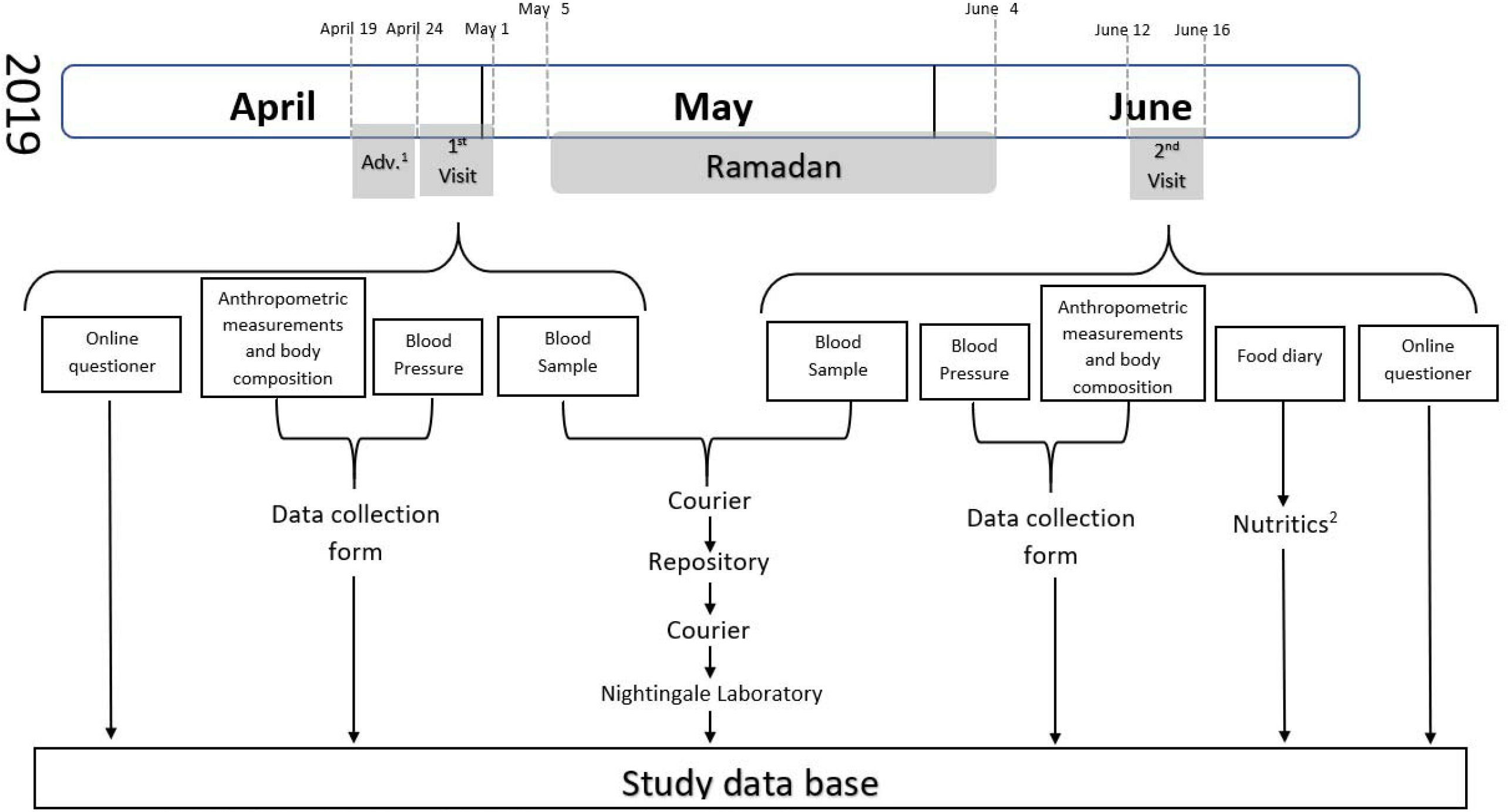
Study design. ^1^ advertising the study in the five mosques. ^2^ a nutrition software.

Clinics were set up from 1 to 5 days in various mosques depending on the expected number of participants. Each clinic was consisted of four stations as follows:

### First station

Initially, participants were asked to read the participant information sheet and sign the informed consent form. Each participant was given a unique barcode. Then, using a tablet and a secure application, participants completed an online questionnaire. The questionnaire included questions on demographics, physical activity, sleep pattern, current health issues, medical history, smoking and alcohol consumption, mental health and fasting history (Appendix 1).

### Second station

The research assistant measured the anthropometric measurements (by calculating the average of two measurements for each parameter) and body composition. Anthropometric measurements include height (using Leicester Height Measure), weight (Marsden digital weighing scale), waist and hip circumference (tape measure). A bioelectrical impedance analyser (Tanita BC-418) was used to measure body composition. Anthropometric measurements and body composition were not collected for participants with pacemakers or metal implants.

### Third station

The phlebotomist measured blood pressure (Omron 705-IT) three times, then collected a blood sample (2 × 9ml EDTA, 2 × 5ml serum separation tubes, 1 X RNA stabiliser). In mosques, blood samples were kept in thermoporters (1 – 6 hours) which were sent within one hour by a courier to the laboratory in Charing Cross Hospital (London, UK) for routine measurements. The remaining samples were aliquoted and stored in a long-term repository (−80 °C).

### Fourth station

At the end, participants were given a food diary to be completed for three days (two weekdays and a weekend day) before Ramadan and three days during Ramadan to observe their diet. The food diary booklet had usage instructions, including an example of food intake for one day completed to show how to describe food or their amount. There was a section dedicated for reporting the participant’s vitamins/minerals or other food supplements.

The food diaries were collected during the second visit.

During the last ten days of Ramadan, we contacted participants using different methods (phone calls, letters and emails) to confirm their appointments after Ramadan and remind them to fill out their food diaries for three days during Ramadan.

After Ramadan, the second visit was done in the same way as the first one. Ethical approval was obtained from the Imperial College Research Ethics Committee (reference: 19IC5138, dated 17/4/2019).

### Food intake

We used Nutritics (nutrition software) to analyse the food diaries (39).

### Metabolomics measurements

A year later, aliquots of plasma were sent via an international courier (https://www.worldcourier.com/) to Nightingale laboratory in Finland for metabolic biomarker profiling (https://nightingalehealth.com/biomarkers).

## Results

Before Ramadan, we collected data from 146 participants, of whom 85 attended the second visit. The group who did not complete the study (n=61) had no daily smokers and was younger than the group who attended the two visits (appendix 2). Of the 85 participants who completed the study, 55 (47.1%) were females. The mean age of the participants was 45.6 ± 15.9 years. Overall, 25.5% of the study population were obese (≥ 30 kg/m^2^), 4.7% were smokers, 14% had diabetes (self-reported), 24% had hypertension (self-reported), and 5.2% had cardiovascular diseases. In addition, 81.2% had fasted the whole month (30 days), and the mean of fasted days during Ramadan was 28.6 days. **Table 1** presents the baseline characteristics of the 85 participants who completed the study. We collected data on anthropometry and blood pressure in both visits for 85 participants (100%), body composition from 84 (98.8%), blood samples from 81 (95.3%) and food diaries from 56 (65.9%) participants. **Figure 3** links types of data collected to locations (mosques).

**Table1:** baseline characteristics of the 85 participants who completed the study by gender.

|  | Men<br>(n=45) | Women<br>(n=40) | Total<br>(n=85) |
| --- | --- | --- | --- |
| **N (%)** | 52.9 | 47.1 | 100 |
| **Age (mean (SD))** | 46.5 (16.3) | 44.6 (15.4) | 45.6 (15.9) |
| 18 – 40 | 33.3 | 30 | 31.8 |
| 40 – 60 | 42.2 | 57.5 | 49.4 |
| 60 – 80 | 22.2 | 12.5 | 17.6 |
| > 80 | 2.2 | 0 | 1.2 |
| **Fasted the whole month (30 days)** | 95.5 | 65 | 81.2 |
| **Ethnicity** |  |  |  |
| Arab | 19 | 13.5 | 16.5 |
| Bangladeshi | 9.5 | 8.1 | 8.9 |
| Indian | 23.8 | 29.7 | 26.6 |
| Pakistani | 4.3 | 18.9 | 16.5 |
| Somali | 19 | 13.5 | 16.5 |
| Other | 14.3 | 16.2 | 15.2 |
| **BMI** |  |  |  |
| < 25 kg/m<sup>2</sup> | 37.8 | 25 | 31.8 |
| ≥ 25 kg/m<sup>2</sup> & < 30 kg/m<sup>2</sup> | 52.2 | 35 | 44.1 |
| ≥ 30 kg/m<sup>2</sup> & < 35 kg/m<sup>2</sup> | 7.7 | 26.2 | 16.5 |
| ≥ 35 kg/m<sup>2</sup> & < 40 kg/m<sup>2</sup> | 2.2 | 11.2 | 6.5 |
| ≥ 40 kg/m<sup>2</sup> | 1.2 | 0 | 2.5 |
| **Marital status** |  |  |  |
| Married/living with a partner | 69 | 78.4 | 73.4 |
| Single | 28.6 | 16.2 | 22.8 |
| Divorced/separated | 2.4 | 5.4 | 3.8 |
| **Chronic diseases** |  |  |  |
| Diabetes | 19 | 8.1 | 14 |
| Hypertension | 26.2 | 21.6 | 24 |
| Cardiovascular diseases | 7.3 | 2.8 | 5.2 |
| **Education** |  |  |  |
| No formal qualification | 7.1 | 18.9 | 12.7 |
| Secondary school or equivalent | 21.4 | 29.7 | 25.3 |
| Higher education: College/HNC/HND | 19 | 24.3 | 21.5 |
| Vocational qualification | 2.3 | 0 | 1.3 |
| Bachelor's degree | 28.5 | 24.3 | 26.6 |
| Postgraduate degree | 21.4 | 2.7 | 12.7 |
| **Smoking** |  |  |  |
| Never | 57.7 | 100 | 77.6 |
| Stopped | 26.6 | 0 | 14.1 |
| Occasionally | 6.6 | 0 | 3.5 |
| Yes, most or all days | 8.8 | 0 | 4.7 |

Percentages are presented for all variables except for age (mean (SD)).

**Figure3:**
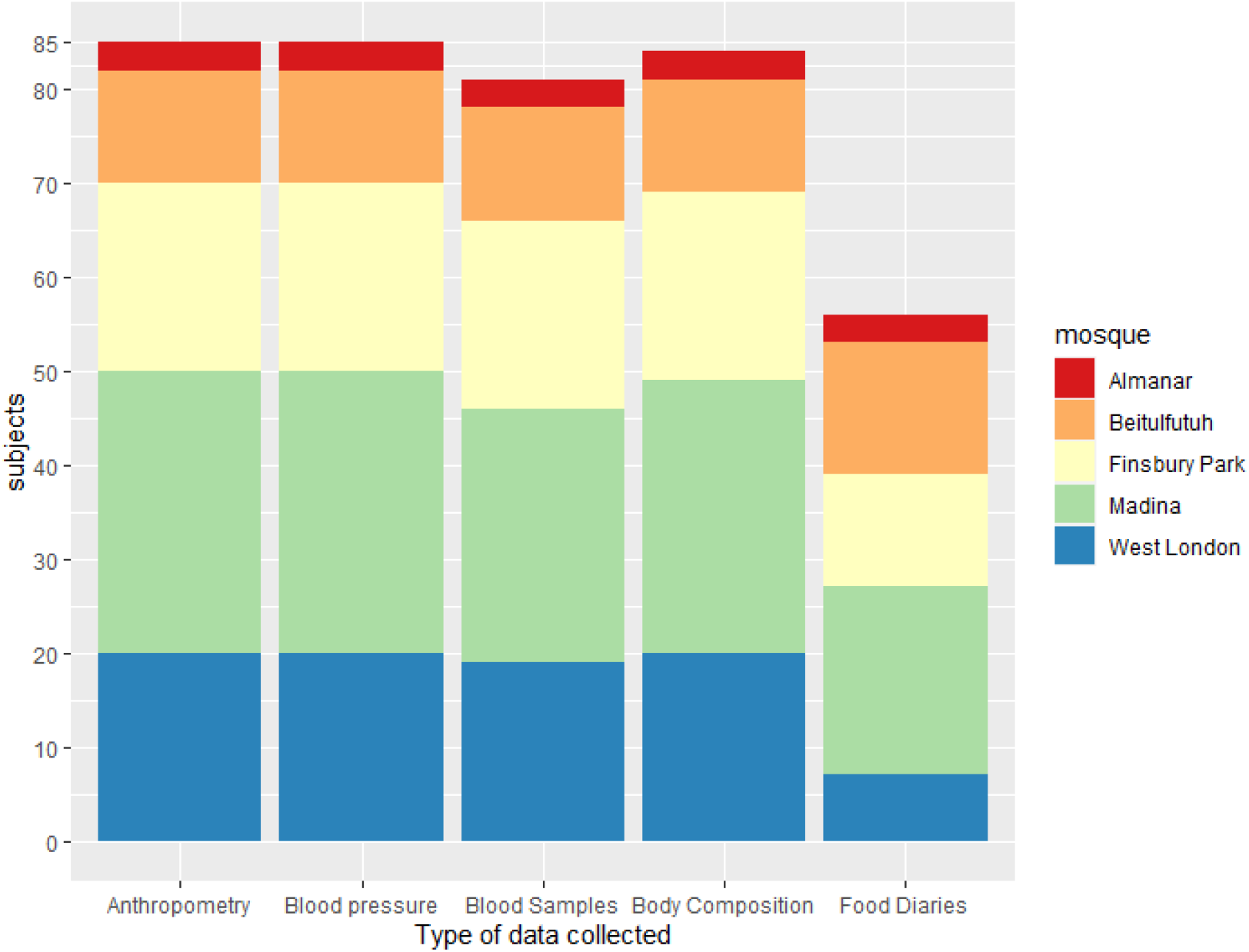
types of collected data by locations (mosques).

After processing raw data of the food diaries, we acquired data on around 190 nutrients, including proteins, lipids, carbohydrates, fibre, vitamins, water and minerals. In addition, Nightingale laboratory applied metabolic biomarker profiling and provided us with data on 249 metabolites for each blood sample (**Fig. 2**).

## Discussion

LORANS recruited a sample from various ethnic groups with different cultures. Collected data included lifestyle, food intake, blood pressure, anthropometry/body composition and metabolic biomarkers. Although the only incentive we offered was receiving personalised feedback on the impact of Ramadan fasting on participants health, people were willing to volunteer and participate. This study has shown that conducting large scale studies in under-studied populations is feasible.

In LORANS, we managed to recruit a sample that is larger than most of the former Ramadan fasting studies. Recruiting individuals in such studies could be challenging, which explains the small samples in most previous studies. Conducting LORANS, we built experience in recruiting large samples with high follow up rates in future. We learned that engaging the mosque administration facilitates approaching and encouraging mosques attendees to participate in the study. Another factor is that all mosques receive a large number of people on Friday midday prayer (Zuhur) compared to other prayers across the weekdays. This massive gathering is an excellent opportunity to spread the word about the study. In addition, door to door advertising in Muslim-majority neighbourhoods would attract more participants. In LORANS, we had limited time to advertise the study since the ethical approval arrived less than two weeks before Ramadan. Many participants plan their holiday for the days after the fasting month. In many cases, they were not available for the particular day that we had the clinic in the mosque for the second visit. Having the clinic for several days in each mosque after Ramadan will increase the number of people who attend the follow up visit.

In summary, LORANS has recruited a multicultural community-based sample from a city with various ethnic groups. The recruited sample was balanced in terms of age groups and gender. Furthermore, LORANS has collected a comprehensive set of data on lifestyle and health which enables us to analyse the data considering all important factors. LORANS further shows that conducting large studies on the health effects of Ramadan fasting is feasible. Engaging the mosque administrations, long-term and extensive advertisement for the study and sufficient time for clinics after Ramadan are essential for recruiting large samples and high rates of follow up.

## Data Availability

All data are available.
There is no reporting checklist available on EQUATOR Network for this manuscript type (, so it is not applicable.

## Acknowledgements

We would like to thank all participants in LORANS. We also thank Ahlam Khamliche, Kimberley Bennett and Jordan Jenkins for their assistance in data collection and management. We thank Aida Abdelwahed, Saredo Said, Rima Mustafa, Hamad Al Jafar, Faisal Al-Ghamdi, Sharmin Akbar, Sadia Zaman, Rahma Hassan, Amna Ahmed, Shifa Bangi, Mahnoor Ahmed, Karim Belhaj, Manal Al Jafar and Yi ZHAO who volunteered to advertise the study and collect data.

## Funding sources

This project was partially funded by the Saudi Embassy in London.

## Notes

### Competing Interest Statement

The authors have declared no competing interest.

### Clinical Trial

This study is not a trail

### Author Declarations

Imperial College Research Ethics Committee

