## Appendix 1 for "London Ramadan Fasting Study (LORANS): Rationale, design, and methods"

### Questionnaire

### Before Ramadan fasting

Barcode

**Personal Information**

**Name:**

**Surname:**

**Address:**

**Postcode:**

**Phone:**

**Mobile:**

**Email address:**

#### Section 1: Demographics and socioeconomic status:

##### 1.1 General Information

**1.1.1 Marital status:**

O Single

O Married/living with a partner

O Divorced/separated

O Widow/widower

**1.1.2 What is your ethnic background?**

O Pakistani

O Indian

O Bangladeshi

O Somali

O Arab

O Turkish

O Persian

O Others (please name):……………………………………

**1.1.3 What is the highest level of education that you have completed?**

O No formal qualification/primary school not completed

O Secondary school or equivalent

O Higher education: College/HNC/HND

O Vocational/ITE (Institute of Technical Education) or equivalent

O Bachelor's degree

O Postgraduate degree

##### 1.2 Employment

**1.2.1 What is your employment status?**

O Unemployed looking for job (Proceed to Section 2)

O Unemployed not looking for job (Proceed to Section 2)

O House wife (Proceed to Section 2)

O Employee (For males, this includes serving full-time National Service)

O Employer (i.e. operate own business or trade with paid work)

O Owns personal business/trade with no employees O Works in family business

O Retired O Student or trainee O Voluntary work

O None of the above

**1.3 Working Pattern and Shift Work**

**1.3.1 In a typical working week, how many hours of paid work (including overtime) do you undertake (not including housewife, childcare, carer)?**

O Less than 10 hours  O 10 to less than 20 hours

O 20 to less than 40 hours  O 40 to less than 50 hours

O 50 to less than 60 hours  O 60 to less than 70 hours

O 70 to less than 80 hours  O 80+ hours

#### Section 2: Physical and Sedentary Activities

**2.1 Thinking of your main occupation, what level of activity is involved?**

Main occupation may also include housewife, part-time work, retiree, voluntary work, etc.

O Sitting most of the time  O Standing most of the time  O Walking most of the time

O Sitting, standing, and walking in equal amounts

O Other work with moderate physical activity (includes moving or lifting objects of moderate weight)

O Physically heavy work (includes moving or lifting heavy objects or activities)

O None of the above

**2.2 Please tell us about the time you have spent per day sitting while using digital devices, during a typical week. For example: watching TV, DVDs, mobile phone, iPad, using a computer etc.**

| Time spent<br>Part of the week | None | Less than one hour per day | Between 1 hour and less than 2 hours per day | Between 2 and 4 hours per day | More than 4 hours per day |
|---|---|---|---|---|---|
| During the weekends | O | O | O | O | O |
| During the weekdays | O | O | O | O | O |

**2.3 Please tell us about the physical activities that you do in a typical week for at least 10 minutes solely for recreation, sport, exercise, or leisure.**

**Indicate the total number of days, and the average amount of time spent in that activity per day.**

| Frequency<br>Activity | Number of days a week | Average time per day |
|---|---|---|
| Walking | | |
| Moderate physical activity<br>(e.g. swimming or bicycling at regular pace, pilates, yoga, doubles tennis, gym) | | |
| Heavy sport activity<br>(e.g. running, fast bicycling, fast swimming, aerobics, weights lifting, singles tennis, football) | | |

**2.4 In the last 4 weeks, which form of transport have you used most often to get about?**

O Car/motor vehicle  O Walk  O Public transport (e.g. MRT, buses, taxis)  O Other

#### Section 3: Sleep Pattern

The following questions relate to your usual sleep habits during the past week. Your answers should indicate the most accurate reply for the majority of days and nights in the past month.

**3.1 When have you usually gone to bed?**
Bed time (hour : minute AM/PM):

**3.2 Approximately how long does it take you to fall asleep at night? (Please give your answer in minutes)**
Number of minutes to fall asleep:

**3.3 When have you usually gotten up in the morning?**
Getting up time (hour : minute AM/PM):

**3.4 How many hours of actual sleep do you get at night? (This may be different than the number of hours you spend in bed)**
Hours of sleep per night:

**3.5 Do you usually sleep in the afternoon? (give the answers according to the last month)**

O Never (proceed to section 4) O Just once or twice in a month O Just a few times in a month O nearly half of the month O Most days or every day

**3.6 How many hours do you usually sleep in the afternoon?**
Hours of sleep per afternoon:

#### Section 4: General and Personal Health

**4.1 In general, how would you rate your overall health?**

O Very good O Good O Fair O Poor O Very poor

#### Section 5: Other Health Issues

**5.1 In the last year, did you have any of the following complains that interfered with your usual activities and last for more than a month?**

| | Never | Just a few times | Once a week | 2 or 3 times per week | Most days |
|---|---|---|---|---|---|
| Headache or migraine | | | | | |
| Poor appetite | | | | | |
| Heartburn | | | | | |
| Abdominal distension or feeling to bloat | | | | | |
| Stomach or abdominal pain | | | | | |
| Constipation | | | | | |
| Neck or shoulder pain | | | | | |
| Back pain | | | | | |
| Hip pain | | | | | |
| Knee pain | | | | | |
| Foot pain | | | | | |
| Stiffness of joints in the morning that lasts more than 30 minutes | | | | | |

#### Section 6: Questions for All Participants

##### 6.1 Diabetes

**6.1.1 Has a doctor ever told you that you have or had diabetes?**

O Yes

O No (Proceed to question 6.2.1)

**6.1.2 How old were you when your diabetes was first diagnosed? (This does not include gestational diabetes)**

Age: O Do not remember

**6.1.3 How is your diabetes being treated? (You can select more than one answer)**

☐ Diet ☐ Increased physical activity ☐ Tablets

☐ Insulin ☐ My diabetes is not being treated

**6.1.4 Did you start insulin within one year of your diagnosis of diabetes?**

O Yes O No O Do not remember O Not applicable

##### 6.2 Cholesterol

**6.2.1 Has a doctor ever told you that you have or had high cholesterol?**

O Yes

O No (Proceed to question 6.3.1)

**6.2.2 How old were you when the doctor first told you that you had high cholesterol?**

Age: O Do not remember

**6.2.3 How is your high cholesterol currently been treated? (You may select more than one answer)**

☐ Diet ☐ Increased physical activity ☐ Tablets ☐ My high cholesterol is not being treated

##### 6.3 High Blood Pressure

**6.3.1 Has a doctor ever told you that you have or had high blood pressure?**

O Yes

O No (Proceed to question 6.4.1)

**6.3.2 How old were you when the doctor first told you that you had high blood pressure?**

Age: O Do not remember

**6.3.3 How is your high blood pressure being treated? (You can select more than one answer)**

☐ Diet ☐ Increased physical activity ☐ Tablets

☐ My high blood pressure is not being treated

##### 6.4 Thyroid function

**6.4.1 Has a doctor ever told you that you have any thyroid problems (hypothyroidism or hyperthyroidism)?**

O Yes - Hyperthyroidism

O Yes - Hypothyroidism

O Yes - I do not know the details

O No (Proceed to section 7)

**6.4.2 How old were you when the doctor first told you that you had thyroid problem?**

Age: O Do not remember

**6.4.3 How is your thyroid problem been treated?**

☐ Yes ☐ No

#### Section 7: Personal Medical History

**Q. Has a doctor ever told you that you have or had any of the following conditions? Indicate the age you were diagnosed and whether or not you received treatment. (You can select more than one answer)**

| Condition | Age of diagnosis | Was/currently being treated (please circle) |
|---|---|---|
| ☐ Heart disease of any causes | | Yes / No |
| ☐ Stroke | | Yes / No |
| ☐ Obesity | | Yes / No |
| ☐ Asthma | | Yes / No |
| ☐ Chronic bronchitis/ emphysema (COPD) | | Yes / No |
| ☐ Allergic rhinitis | | Yes / No |
| ☐ Any seizure disorders including epilepsy (seizure within the last 3 years and currently in antiepileptic medication within the last year) | | Yes / No |
| ☐ Severe depression | | Yes / No |
| ☐ Bipolar Disorder | | Yes / No |
| ☐ Anxiety | | Yes / No |
| ☐ Glaucoma | | |
| ☐ Kidney/Bladder stone | | |
| ☐ Chronic kidney disease of any causes | | |
| ☐ Rheumatoid arthritis | | |
| ☐ Osteoarthritis | | Yes / No |
| ☐ Osteoporosis | | Yes / No |
| ☐ Other disease: ______________________________ | | Yes / No |
| ☐ None of the above | | |

#### Section 8: Housing Information

**8.1 What is the total monthly income for your household?**

Options will be added later

#### Section 9: Smoking

##### 9.1 Smoking Habits

**9.1.1 Do you currently smoke tobacco?**

O No, have never smoked (proceed to section 10)

O No, just have tried once or twice (proceed to section 10)

O No, stopped smoking (proceed to question 9.3.1)

O Yes, only occasionally (proceed to question 9.2.1)

O Yes, on most or all days (proceed to question 9.2.1)

##### 9.2 Current Smokers

**9.2.1 What do usually smoke?** (you can select more than one answer)

O Cigarettes O Shisha (Pipe) O Cigars O Bidi (hand rolled cigarettes) O None of the above

**9.2.2 How often do you smoke?**

| Smoking Type | Frequency per day |
|---|---|
| Cigarettes / Beedi (hand rolled cigarettes) | …… Cigarettes per day |
| Cigars | …… Cigars per day |
| Shisha / Pipe / Hookah | …… Times per day |

**9.2.3 How old were you when you first started smoking?**

Age: O Do not remember

**9.2.4 How many years have you smoked so far in total?**

….. Years

##### 9.3 Past smokers

**9.3.1 What did you usually smoke? (you select more than one answer)**

O Cigarettes O Shisha (Pipe) O Cigars O Bidi (hand rolled cigarettes) O None of the above

**9.3.2 How many cigarettes did you typically smoke each day?**

| Smoking Type | Frequency per day |
|---|---|
| Cigarettes / Beedi (hand rolled cigarettes) | …… Cigarettes per day |
| Cigars | …… Cigars per day |
| Shisha / Pipe / Hookah | …… Times per day |

**9.3.3 How old were you when you first started smoking?**

Age: O Do not remember

**9.3.4 How old were you when you stopped smoking?**

Age: O Do not remember

**9.3.5 How many years have you smoked so far in total?**

….. Years

#### Section 10: Current Drinking Habits

**10.1 How often did you drink alcohol during the last month?**

O Never O Special occasions only (less than once per month)

O One to three times a month O Once or twice a week

O Three or four times a week O Daily or almost daily

#### Section 11: Feelings and Mood

| Over the **last month**, how often have you been bothered by any of the following problems? | | | | |
|---|---|---|---|---|
| | Not at all | Several days | More than half the days | Nearly everyday |
| a. Little interest or pleasure in doing things | O | O | O | O |
| b. Feeling down, depressed, or hopeless | O | O | O | O |
| c. Trouble falling or staying asleep, or sleeping too much | O | O | O | O |
| d. Feeling tired or having little energy | O | O | O | O |
| e. Poor appetite or overeating | O | O | O | O |
| f. Feeling bad about yourself, or that you are a failure, or have let yourself or your family down | O | O | O | O |
| g. Trouble concentrating on things, such as reading the newspaper or watching television | O | O | O | O |
| h. Moving or speaking so slowly that other people could have noticed? Or the opposite – being so fidgety or restless that you have been moving around a lot more than usual | O | O | O | O |
| i. Thoughts that you would be better off dead or hurting yourself in some way | O | O | O | O |

#### Section 12: Fasting History

**Do you fast every year?**

O Yes O No

**Did you fast during Ramadan last year?**

O Yes O No

**How many days did you fast during Ramadan last year?**

O the whole month O more than 20 days O more than 10 days O less than 10 days

### Questionnaire

### After Ramadan fasting

**Personal Information**

**Name:**

**Surname:**

**Address:**

**Postcode:**

**Phone:**

**Mobile:**

**Email address:**

#### Section 1: Demographics and socioeconomic status:

##### 1.1 Working Pattern and Shift Work

**1.1.1 In a typical working week <u>during Ramadan</u>, how many hours of paid work (including overtime) do/did you undertake (not including housewife, childcare, carer)?**

O Less than 10 hours  O 10 to less than 20 hours

O 20 to less than 40 hours  O 40 to less than 50 hours

O 50 to less than 60 hours  O 60 to less than 70 hours

O 70 to less than 80 hours  O 80+ hours

#### Section 2: Physical and Sedentary Activities

**2.1 What level of activity was involved with your work <u>during Ramadan</u>?**

Main occupation may include <u>housewife</u>, part-time work, retiree, voluntary work, etc.

O Sitting most of the time  O Standing most of the time  O Walking most of the time

O Sitting, standing, and walking in equal amounts

O Other work with moderate physical activity (includes moving or lifting objects of moderate weight)

O Physically heavy work (includes moving or lifting heavy objects or activities)

O None of the above

**2.2 Please tell us about the time you have spent sitting per day while using digital devices, in a typical week <u>during Ramadan</u>. For example: watching TV, DVDs, mobile phone, iPad, using a computer etc.**

| Time spent<br>Part of<br>The week | None | Less than one hour per day | Between 1 hour and less than 2 hours per day | Between 2 and 4 hours per day | More than 4 hours per day |
|---|---|---|---|---|---|
| During the weekends | O | O | O | O | O |
| During the weekdays | O | O | O | O | O |

**2.3 Please tell us about the physical activities that you did in a typical week <u>during Ramadan</u> for at least 10 minutes solely for recreation, sport, exercise, or leisure.**
**Indicate the total number of days, and the average amount of time spent in that activity per day.**

| Frequency<br>Activity | Number of days a week | Average time per day |
|---|---|---|
| Walking | | |
| Moderate physical activity<br>(e.g. swimming or bicycling at regular pace, pilates, yoga, doubles tennis, gym) | | |
| Heavy sport activity<br>(e.g. running, fast bicycling, fast swimming, aerobics, weights lifting, singles tennis, football) | | |

**2.4 <u>During Ramadan</u>, which form of transport have you used most often to get about?**

O Car/motor vehicle  O Walk  O Public transport (e.g. MRT, buses, taxis)  O Other

#### Section 3: Sleep Pattern

The following questions relate to your usual sleep habits during **Ramadan**. Your answers should indicate the most accurate reply for the majority of days and nights in **Ramadan**.

**3.1 When have you usually gone to bed?**
Bed time (hour : minute AM/PM):

**3.2 Approximately how long did it take you to fall asleep at night? (Please give your answer in minutes)**
Number of minutes to fall asleep:

**3.3 Did you get up at down for dawn meal (Sahur/Sehri) during Ramadan?**

O Yes O No (proceed to question 3.7)

**3.4 At what time did you get up for dawn meal (Sahur/Sehri)?**
Getting up time (hour : minute AM):

**3.5 Were you going back to sleep after dawn meal?**

O Yes O No (proceed to question 3.8)

**3.6 At what time do you go to bed again after dawn meal?**
Bed time (hour: minute AM):

**3.7 When have you usually gotten up during Ramadan?**
Getting up time (hour : minute AM/PM):

**3.8 How many hours of actual sleep did you get at night during Ramadan? (This may be different than the number of hours you spend in bed)**
Hours of sleep per night:

**3.9 Did you usually sleep in the afternoon during Ramadan?**

O Never (proceed to section 4) O Just for a few days (1-5) O nearly half of the month O Most days or every day

**3.10 How many hours did you usually sleep in the afternoon?**
Hours of sleep per afternoon:

#### Section 4: General and Personal Health

**4.1 In general, how would you rate your overall health during Ramadan?**

O Very good O Good O Fair O Poor O Very poor

#### Section 5: Other Health Issues

**5.1 Did you have any of the following complains that interfered with your usual activities during Ramadan?**

| | Never | Just a few times | Once a week | 2 or 3 times per week | Most days |
|---|---|---|---|---|---|
| Headache or migraine | | | | | |
| Poor appetite | | | | | |
| Heartburn | | | | | |
| Abdominal distension or feeling to bloat | | | | | |

| |
|---|
| Stomach or abdominal pain |
| Constipation |
| Neck or shoulder pain |
| Back pain |
| Hip pain |
| Knee pain |
| Foot pain |
| Stiffness of joints in the morning that lasts more than 30 minutes |

**5.2 Were you newly diagnosed with a disease during the month of Ramadan?**

O Diabetes  O Hypertension  O High cholesterol  O Other (name it)  O None

#### Section 5: Smoking

**5.1 Smoking Habits**

**5.1.1 Were you regularly smoking tobacco before or during Ramadan?**

o Yes  o No (Proceed to section 6)

**5.1.2 What did you usually smoke during Ramadan?** (you select more than one answer)

O Cigarettes  O Shisha (Pipe)  O Cigars  O Bidi (hand rolled cigarettes)  O None of the above

**5.1.3 How often did you smoke on average during Ramadan?**

| Smoking Type | Frequency per day |
|---|---|
| Cigarettes / Beedi (hand rolled cigarettes) | …… Cigarettes per day |
| Cigars | …… Cigars per day |
| Shisha / Pipe / Hookah | …… Times per day |

#### Section 6: Drinking Habits

**6.1 Did you drink alcohol during Ramadan?**

O Never  O Only once

O two or three times  O Once or twice a week

O Three or four times a week  O Daily or almost daily

#### Section 7: Feelings and Mood

How often have you been bothered by any of the following problems **during Ramadan**?

| | Not at all | Several days | More than half the days | Nearly everyday |
|---|---|---|---|---|
| a. Little interest or pleasure in doing things | O | O | O | O |
| b. Feeling down, depressed, or hopeless | O | O | O | O |
| c. Trouble falling or staying asleep, or sleeping too much | O | O | O | O |
| d. Feeling tired or having little energy | O | O | O | O |
| e. Poor appetite or overeating | O | O | O | O |
| f. Feeling bad about yourself, or that you are a failure, or have let yourself or your family down | O | O | O | O |

| | | | | |
|---|---|---|---|---|
| g. Trouble concentrating on things, such as reading the newspaper or watching television | O | O | O | O |
| h. Moving or speaking so slowly that other people could have noticed? Or the opposite – being so fidgety or restless that you have been moving around a lot more than usual | O | O | O | O |
| i. Thoughts that you would be better off dead or hurting yourself in some way | O | O | O | O |

#### Section 8 - Fasting

**Did you fast the whole month of Ramadan this year?**

o Yes (proceed to section 1)  o No ( if No, please indicate how many days) ……………. Days

**What was the reason for not fasting?**

O Sickness  o travelling  o period (females)

O other reason ………………………..
