## Appendix 2 for "London Ramadan Fasting Study (LORANS): Rationale, design, and methods"

Appendix 2: A comparison between participants who completed the study to those who dropped out.

| Variable | Sub-groups | Study participants (n=85) | Individuals who didn't attend the second visit (n=61) | p-value |
|---|---|---|---|---|
| **Age (mean ± SD)** | Total | 45.6 ± 15.9 | 39 ± 21.3 † | 0.002 |
| | 18 – 40 years (%) | 31.8% | 55.7% | |
| | 40 – 60 years (%) | 49.4% | 39.3% | |
| | 60 – 80 years (%) | 17.6% | 11.5% | |
| | > 80 years (%) | 1.2% | 3.3% | |
| **Gender (male %)** | | 52.9% | 49.2 % | 0.80 |
| **Ethnic background (%)** | Pakistani | 16.5% | 18.8% | 0.36 |
| | Indian | 26.6% | 35.4% | |
| | Bangladeshi | 8.9% | 6.3% | |
| | Somali | 16.5% | 22.4% | |
| | Arab | 16.5% | 4.2% | |
| | Other | 15.2% | 12.5% | |
| **Marital status (%)** | Single | 22.8% | 32% | 0.49 |
| | Married/living with a partner | 73.4 % | 66% | |
| | Divorced/separated | 3.8 % | 2.1% | |
| **With Chronic diseases (%)** | Diabetes | 25.9% | 13.1% | 0.83 |
| | Hypertension | 44.7% | 13.1% | 0.48 |
| | Cardiovascular diseases | 9.4% | 6.6% | 0.42 |
| **Education (%)** | No formal qualification | 12.7% | 10.6% | 0.31 |
| | Secondary school or equivalent | 25.3% | 19.1% | |
| | Higher education: College/HNC/HND | 21.5% | 14.9% | |
| | Vocational qualification | 1.3% | 8.5% | |
| | Bachelor's degree | 26.6% | 27.7% | |
| | Postgraduate degree | 12.7% | 19.1% | |
| **Smoking (%)** | Never | 77.6% | 91.5% | 0.02 |
| | Stopped | 14.1% | 2.1% | |
| | Occasionally | 3.5% | 6.4% | |
| | Yes, most or all days | 4.7% | 0% | |

† median and interquartile range
